# Agentic memory-augmented retrieval and evidence grounding for medical question-answering tasks

**DOI:** 10.1101/2025.08.06.25333160

**Authors:** Shuyue Jia, Subhrangshu Bit, Varuna H. Jasodanand, Yi Liu, Vijaya B. Kolachalama

## Abstract

**Objective:** To evaluate if a tool-using agent-based system utilizing large language models (LLMs) for medical question-answering (QA) tasks outperforms standalone LLMs.

**Methods:** We developed a unified, open-source LLM-based agentic system that integrates document retrieval, re-ranking, evidence grounding, and diagnosis generation to support dynamic, multi-step medical reasoning. Our system features a lightweight retrieval-augmented generation pipeline coupled with a cache-and-prune memory bank, enabling efficient long-context inference beyond standard LLM limits. The system autonomously invokes specialized tools, eliminating the need for manual prompt engineering or brittle multi-stage templates. We compared the agentic system against standalone LLMs on various medical QA benchmarks.

**Results:** Evaluated on five well-known medical QA benchmarks, our system outperforms or closely matches state-of-the-art proprietary and open-source medical LLMs in multiple-choice and open-ended formats. Specifically, our system achieved accuracies of 82.98% on USMLE Step 1 and 86.24% on USMLE Step 2, surpassing GPT-4’s 80.67% and 81.67%, respectively, while closely matching on USMLE Step 3 (88.52% vs. 89.78%).

**Conclusion:** Our findings highlight the value of combining tool-augmented and evidence-grounded reasoning strategies to build reliable and scalable medical AI systems.

## INTRODUCTION

Large language models (LLMs) are transforming medical research and practice, showing promise in tasks such as medical question answering (QA) and clinical decision support ^1–4^. However, several challenges continue to limit their reliability and scalability in real-world applications. One major concern is hallucination, which involves the generation of confident yet factually incorrect or ungrounded responses. Another issue is the limited context window of current LLMs, which restricts the amount of information they can process at once, often necessitating retrieval-augmented generation (RAG) pipelines. While RAG improves grounding, it typically incorporates only a subset of relevant evidence, which can introduce bias or lead to incomplete assessments ^5–7^. Additionally, many diagnostic systems require manually engineered multi-stage prompts^8–11^, making them difficult to scale and adapt.

To improve reliability, recent work has explored continual pretraining on medical corpora ^12–14^, instruction fine-tuning and reinforcement learning to enhance medical reasoning ^12,14–16^, and RAG frameworks for grounding model outputs in high-quality evidence ^5,6,8,9^. Despite this progress, most systems focus on either improving reasoning or grounding, rather than jointly optimizing both. Yet, evidence-based medical practice requires sound diagnostic reasoning and alignment with high-quality clinical evidence ^17^. Recent advances in medical reasoning and diagnosis using LLMs have generally progressed along three major directions. The first focuses on continual pretraining of publicly available general-purpose LLMs on domain-specific medical corpora, including textbooks, research articles, and podcast transcripts ^12–14,18^. The second emphasizes instruction tuning or reinforcement learning using medical datasets, which may be manually curated or generated using systems like ChatGPT; these models are fine-tuned through supervised learning or reward feedback to improve chain-of-thought reasoning and emulate realistic doctor-patient interactions ^12,14–16^. Both strategies aim to enhance the medical reasoning skills of general-purpose LLMs, yet despite gains on benchmarks, these models remain vulnerable to hallucinating factually incorrect or unsupported content. A third direction has explored RAG pipelines to address hallucination risks by grounding model outputs in retrieved medical documents ^5,6,8,9,11^, improving factuality but often prioritizing retrieval without simultaneously optimizing for complex diagnostic reasoning. These observations motivate the need for unified approaches that seamlessly combine robust evidence retrieval with dynamic, multi-step medical reasoning.

Medical AI agents leverage the reasoning and language capabilities of LLMs to perform complex clinical tasks, including diagnosis and decision support ^19^. Recent work on medical AI agents has evolved in three directions. The first focuses on role simulation, where agents emulate clinical roles such as doctors, nurses, and patients in simulated environments; these multi-agent systems aim to model clinical workflows through collaborative interactions and reasoning ^20–24^. The second centers on visual question answering, where agents are augmented with domain-specific tools, such as segmentation models for identifying salient regions in medical images and optical character recognition systems for processing textual content from clinical documents ^25,26^. While promising, these approaches often lack explicit mechanisms for diagnostic reasoning or robust integration with large-scale medical knowledge bases. The third involves tool-augmented LLMs, where agents are equipped with capabilities such as document retrieval, function calling, and database access; however, these systems often depend on resource-intensive model retraining or rely on closed-source, paid platforms (e.g., GPT-4) ^2,27–30^, limiting scalability and transparency. Current trends point toward an unmet need for flexible, lightweight, and interpretable frameworks that can dynamically orchestrate evidence gathering, reasoning, and clinical decision-making without prohibitive computational overhead. Our work addresses this emerging need by designing a modular, open, and deployment-friendly system for medical diagnosis support.

To address these challenges, we present a unified, agentic system that integrates evidence retrieval, reranking, grounding, and diagnosis generation. Our system uses open-source tools to orchestrate the entire pipeline, from query analysis to final diagnosis, drawing from a comprehensive evidence base that includes PubMed abstracts and full texts, ClinicalTrials.gov entries, the *New England Journal of Medicine* (NEJM) case reports, medical textbooks, and curated Wikipedia content ^5,31–34^. To efficiently manage this information, we adopted a two-stage retrieval process including coarse-grained retrieval followed by fine-grained reranking. To circumvent the limitations of LLM context windows, we introduced a cache-and-prune memory mechanism that retains high-relevance documents across reasoning steps, allowing the system to make informed decisions over extended sequences. Our contributions are summarized as follows: (i) We propose a unified, fully-automated system that integrates document retrieval and reranking, evidence grounding, and diagnosis generation through an open-source AI agent; (ii) We present a tool-augmented LLM-based agentic architecture that enables dynamic multi-step tool use, eliminating the need for manually engineered prompts or multi-stage pipelines; (iii) We introduce a cache-and-prune memory bank mechanism that efficiently extends the retention of relevant documents for evidence grounding, enhancing diagnostic accuracy and computational efficiency.

## METHODS

Our agentic system comprises three core components (Fig. 1): (1) a lightweight RAG pipeline for efficient evidence retrieval and reranking; (2) an open-source LLM-based agent that autonomously orchestrates diagnostic workflows, from retrieval to reasoning, grounding, and diagnosis generation; and (3) a cache-and-prune memory bank that preserves relevant long-context documents to improve evidence use and diagnostic accuracy. Below we provide additional details on these components.

**Figure 1.**
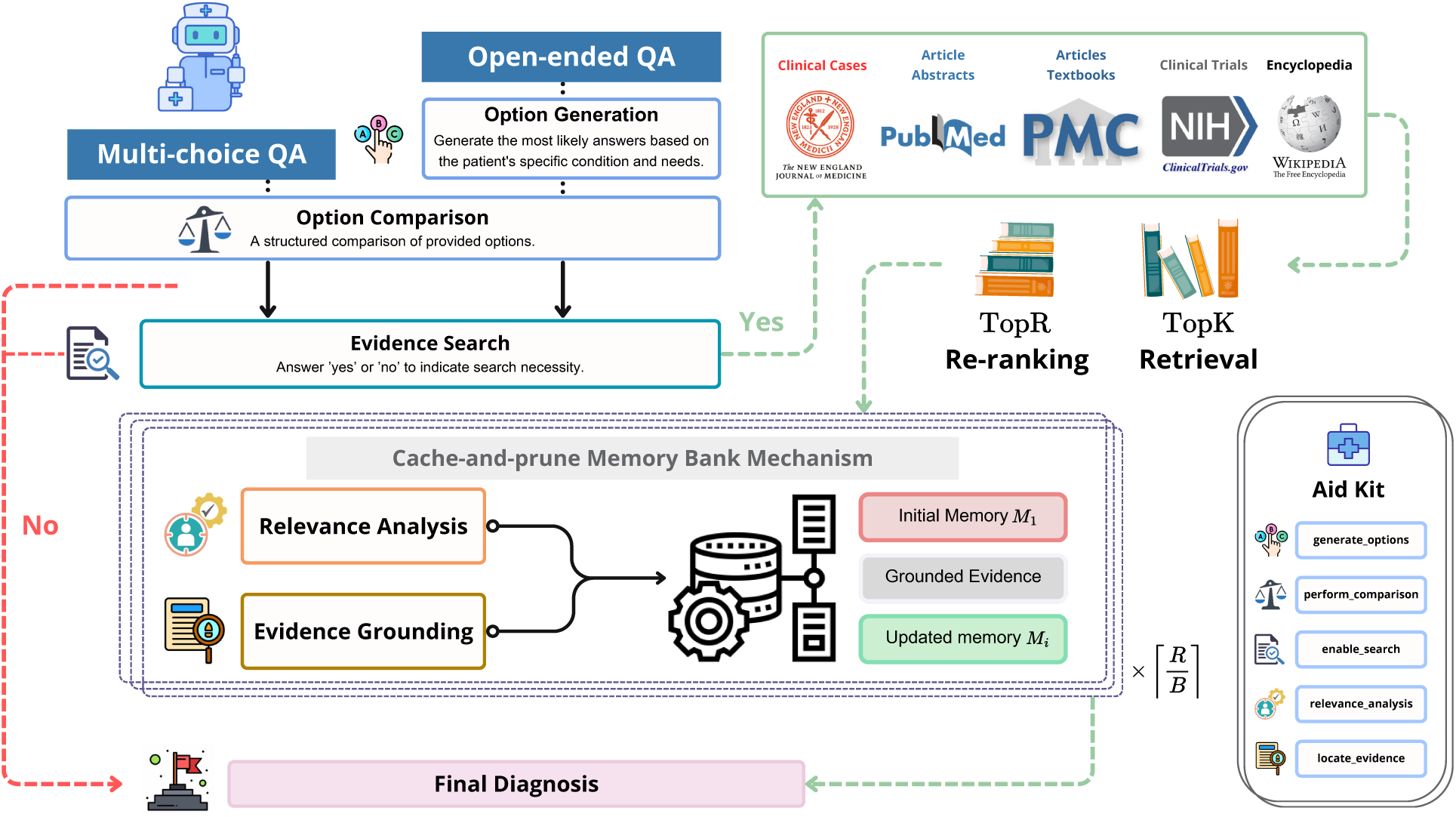
Overview of the agentic system. Our pipeline is powered by an open-source LLM-based agent that operates within a fully automated, dynamic workflow. When presented with either multiple-choice or open-ended medical questions, the agent leverages a suite of specialized tools to generate a structured comparison of answer choices or to synthesize plausible options in open-ended scenarios. It then dynamically assesses whether external evidence is needed to answer the question. If no external information is required, the agent proceeds directly to produce a final diagnosis. Otherwise, it initiates a retrieval process, querying a curated knowledge base to obtain the TopK relevant documents and rerank the TopR most informative sources. This evidence pool includes clinical case reports from NEJM, article abstracts from PubMed, full-text articles and textbooks from PubMed Central, clinical trials from ClinicalTrials.gov, and general content from Wikipedia. To manage long-context documents efficiently, the agent employs a cache-and-prune memory bank mechanism. It iteratively reviews *B* documents in ⌈*R/B*⌉ batches until sufficient information is gathered, ensuring optimal comprehension within the model’s context window. After synthesizing the selected evidence, the agent integrates key insights to deliver a grounded diagnosis. Its performance is further enhanced by an aid kit of five custom-designed tools, detailed in Supplementary Section Designed tools.

### Lightweight RAG pipeline

We implemented a lightweight yet effective RAG pipeline to acquire relevant medical evidence tailored to patient-specific queries. This pipeline consists of two main stages: document retrieval and evidence reranking. In the retrieval stage, we utilized `SPECTER`, a semantic retriever trained with citation-informed objectives, which improved document-level representation, making it particularly effective in biomedical and scientific domains ^5,35^. Denoted as *ϕ*, `SPECTER` retrieves documents by computing semantic similarity between the query representation **x** and document embeddings from the evidence corpus *V*, using L2 distance as the similarity metric:

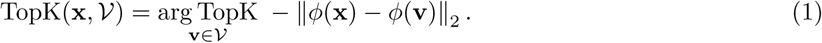

As summarized in Table 1, our evidence corpus includes diverse resources such as research paper abstracts and full texts, medical textbooks, clinical case reports, clinical trials, and curated Wikipedia articles. These are drawn from publicly accessible databases such as PubMed, PubMed Central, ClinicalTrials.gov, and Wikipedia. To refine the quality of retrieved TopK evidence, we implemented a reranking stage. Here, a quantized general text embedding model, `gte-Qwen2-7B-instruct`, was used to score and rank the candidate snippets at a finer granularity, and denoted as *ψ* ^36,37^. This ensures that the top-ranked documents are semantically aligned with the query and optimally suited for downstream diagnostic reasoning:

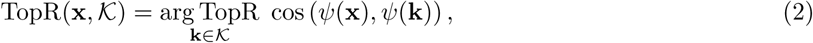

**Table 1.** Performance evaluation on multiple choice medical QA benchmarks. Accuracy scores across five benchmarks: USMLE Step 1–3, MedQA, and MedExpQA. The table compares our agentic system with proprietary (GPT-4, ChatGPT) and open-source (BioMistral, OpenBioLLM, UltraMedical, PodGPT) language models. **Bold** and <u>underlined</u> values denote the best and second-best performances for each benchmark, respectively.

| Model | USMLE Step 1 | USMLE Step 2 | USMLE Step 3 | MedQA | MedExpQA |
| --- | --- | --- | --- | --- | --- |
| GPT-4 | <u>80.67</u> | <u>81.67</u> | **89.78** | **78.87** | N/A |
| ChatGPT | <u>51.26</u> | <u>60.83</u> | 58.39 | 50.82 | N/A |
| BioMistral (7B) | 34.04 | 37.61 | 37.70 | 41.01 | 37.60 |
| OpenBioLLM (8B) | 47.87 | 44.04 | 50.00 | 47.84 | 43.20 |
| UltraMedical (8B) | 42.55 | 27.52 | 34.43 | 38.49 | 35.20 |
| OpenBioLLM (70B) | 69.15 | 70.64 | 68.85 | 69.13 | <u>71.20</u> |
| UltraMedical (70B) | 70.21 | 55.05 | 56.56 | 52.32 | 50.40 |
| PodGPT (70B) | 73.40 | 72.48 | 74.59 | 65.04 | 63.20 |
| **Ours** | **82.98** | **86.24** | <u>88.52</u> | <u>73.29</u> | **78.40** |

where *K* represents the pool of documents retrieved from the six data sources, and *R* denotes the final ranked subset selected for use by the AI agent. Together, these two stages ensured that only the most relevant, high-quality evidence is forwarded for diagnostic processing. This design mitigates hallucination risks and supports accurate, grounded medical reasoning.

### Agent for diagnostic workflow

We integrated an open-source LLM-based agent *π* as the core multi-step reasoning engine of our system to enable autonomous and interpretable medical decision-making. This agent orchestrates the entire diagnostic workflow, including document retrieval and reranking, patient query interpretation, evidence grounding, and diagnosis generation. We designed the agent to operate using a set of predefined tools (See Section), eliminating the need for manually crafted prompts or rigid, hard-coded stages. Each tool encapsulated a specific function, such as querying external evidence sources, grounding highly-relevant documents, or synthesizing diagnostic conclusions. This allows the agent to perform complex clinical tasks in a structured and interpretable manner. By leveraging explicit tool usage and structured reasoning, the agent interacted dynamically and efficiently with the RAG pipeline and memory bank, enabling long-context, evidence-based clinical inference.

Specifically, in the initial step, given a predefined set of tools *T*, the patient’s background and medical query *Q*, and instructions *I*, the AI agent generates a response sequence **y** following an autoregressive policy:

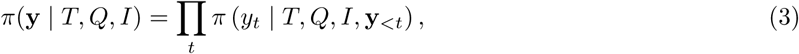

where **y***_<t_* denotes the previously generated tokens up to time step *t* − 1.

Furthermore, at each step of the multi-step reasoning process, the agent autonomously selects the most appropriate tool to address the current subtask and produces intermediate responses in a multi-turn conversational format. Let *C* denote the full conversation history. At each step, the agent selects an action *a* from the action space *A*. Formally,

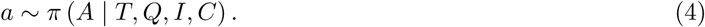

During execution, each intermediate reasoning step produced by the agent, along with any corresponding tool outputs, is appended to the conversation history *C*, enabling coherent multi-turn interactions. This modular tool-based design empowers the agent to flexibly respond to a wide range of clinical queries while ensuring transparency, reproducibility, and traceability throughout the diagnostic workflow. A detailed description of each tool’s output parameters is provided in Fig. 4. Unlike traditional prompt engineering approaches, the agent autonomously determines when and how to invoke each tool through multi-step reasoning. This enables transparent, step-by-step justification of clinical decisions grounded in retrieved evidence. Importantly, the entire workflow operates locally, preserving patient privacy and minimizing reliance on proprietary APIs or cloud-based infrastructure.

### Cache-and-prune memory bank mechanism

To overcome the context window limitations of LLMs and ensure persistent access to relevant evidence for the final diagnostic response, we implemented a cache-and-prune memory bank mechanism. This memory module functions as an external, dynamically updated storage that retains high-relevance documents retrieved and reranked during earlier stages of the pipeline. As shown in Algorithm 1, at each reasoning step indexed by *i*, the AI agent stores the grounded evidence in the memory bank *M_i_*. During the final diagnosis generation, the agent accesses *M_i_*, enabling long-horizon reasoning across multi-turn interactions. To avoid information overload, we designed a cache-and-prune mechanism that filters out outdated or unused evidence, guided by grounding tool usage patterns:

#### Algorithm 1 Agentic memory-augmented retrieval and evidence grounding system

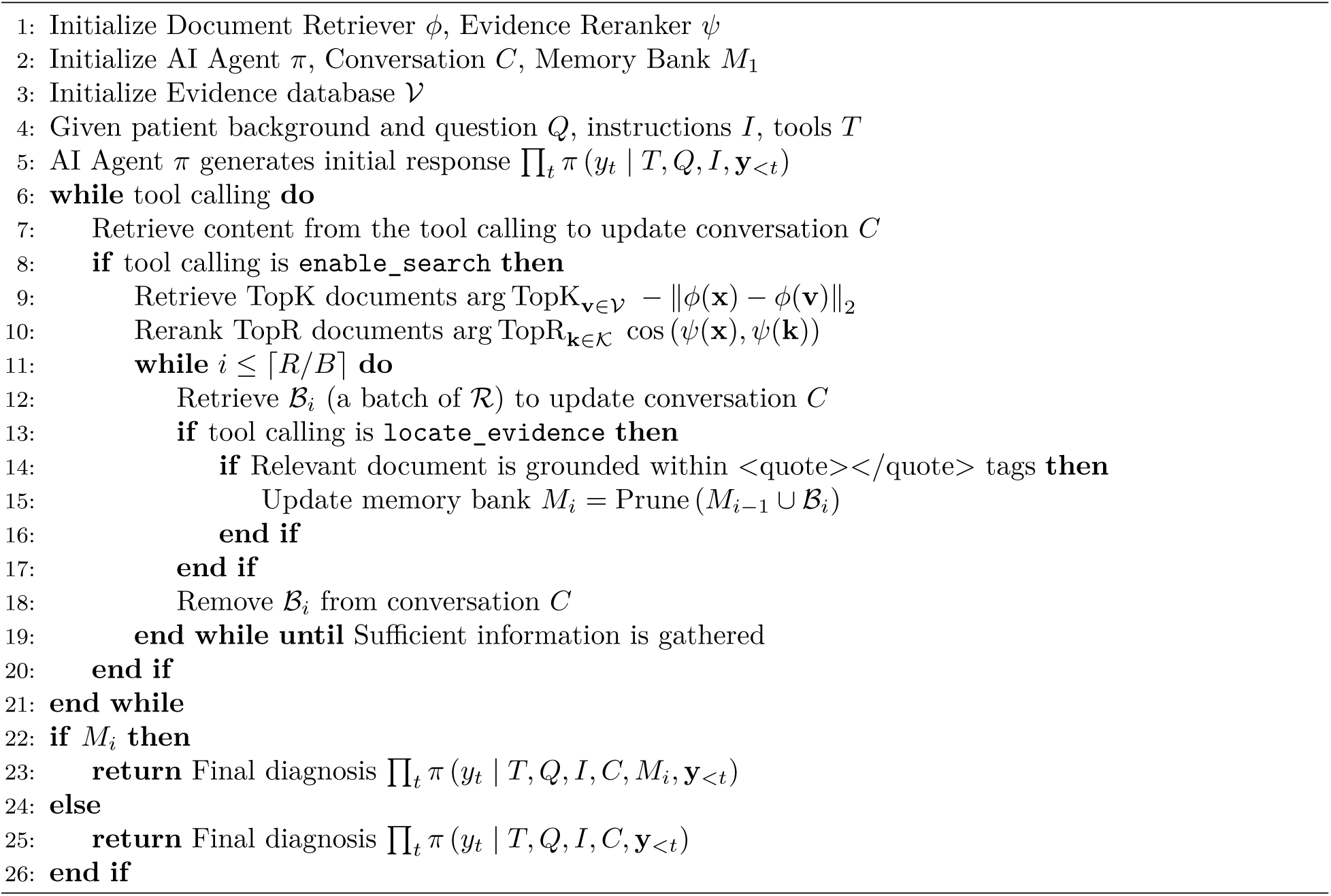

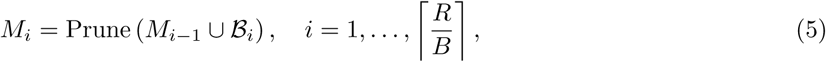

where 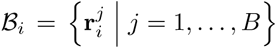 represents the top-ranked documents from each reranked batch *R*, and Prune(·) is a logistic filtering function that removes documents that are not grounded by the AI agent. The final diagnosis is synthesized by conditioning on the complete conversational context, task, instructions, and the curated memory bank *M_i_*:

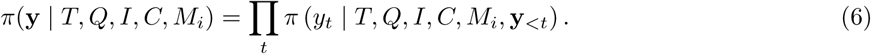

Unlike standard RAG pipelines, which statically inject evidence into the prompt and risk truncation, our memory bank enables selective retention of key information and strategic pruning of less relevant content. This design supports broader context integration and sustained reasoning, mitigating fixed-window constraints and ensuring that only the most salient knowledge informs the agent’s output ^5^.

### Implementation details

All experiments were conducted locally on a distributed setup with four NVIDIA L40S GPUs, powered by the vLLM inference engine ^38^. We employed `Qwen2.5-72B-Instruct` as the primary backbone (i.e., AI agent), with the tensor parallelism and pipeline parallelism settings configured to 4 and 1, respectively. By default, the sampling parameters were set to a temperature of 0 and top_p of 1. To address occasional issues with final answer extraction, we re-evaluated the experiments with a temperature of 0.7 and top_p of 0.8. Due to diminished instruction following capabilities after enabling the static YaRN technique, we assigned the maximum context window to 32,768 tokens ^39^. In practice, however, we observed an effective context window limit of approximately 10,000 tokens. For each multi-turn conversation, we restricted the maximum number of tokens to 8, 192. Additionally, we selected the top 3 most relevant evidence documents for the baseline model that operates without tool access. For evidence retrieval, we fixed TopK = 32 per source, resulting in 192 candidate documents from six sources. After reranking, we selected TopR = 32 documents for use by the agent in downstream tasks ^5,6^. Lastly, the cache-and-prune memory bank operates with a default batch size *B* = 4 for incremental evidence integration and pruning.

## EXPERIMENTAL SETTINGS

### Database for evidence retrieval

To ensure grounding in credible and up-to-date medical evidence, we assembled a comprehensive evidence corpus drawn from six trusted sources. They include peer-reviewed articles from PubMed Central, medical textbooks curated from the NLM LitArch Open Access Subset, and registered clinical trials from the National Library of Medicine at the U.S. National Institutes of Health ^31,32,34^. To enhance clinical relevance and provide real-world diagnostic context, we also incorporated clinical case reports published since 2016 in NEJM ^33^. We also included two supplementary sources, article abstracts and Wikipedia entries, originally curated by Xiong et al. ^5^. Section Database for evidence retrieval in the supplementary materials includes a detailed summary and description of each source included in our evidence retrieval database.

### Benchmark evaluation across question formats

To evaluate the performance of our agentic system, we used five widely adopted medical question answering benchmarks: the United States Medical Licensing Examination (USMLE) Step 1, Step 2, and Step 3, and the English subsets of MedQA and MedExpQA ^6,40,41^. These datasets encompass a range of medical knowledge, clinical reasoning, and decision-making skills, and are well-established standards for evaluating LLMs. See Supplementary Section Experimental benchmarks and Table 3 for more details.

We ran experiments in two settings to test our approach: (1) multiple-choice QA, where models choose from given answer options, and (2) open-ended QA, where models generate answers without being given choices. We compared the performance of the agent against proprietary and open-source medical LLMs. Proprietary models included OpenAI GPT-4 and GPT-3.5 (i.e., ChatGPT), while the open-source models evaluated were BioMistral (7B), OpenBioLLM (8B/70B), UltraMedical (8B/70B), and PodGPT (70B) ^2,18,42–44^. We provided a detailed description of these models and our used prompts in Supplementary Section Backbone large language models and Used prompts. To ensure a fair comparison, we manually ran all open-source models using the vLLM serving engine and applied a consistent zero-shot direct-response prompt. This decision was based on our observation that the performance of some models tended to degrade when presented with more complex instruction prompts. We also set model-specific maximum input lengths and generation token limits to accommodate varying context window sizes. See Supplementary Section Database for evidence retrieval for more details. The full source code developed for this study, including all implementation and evaluation scripts, will be made publicly available on GitHub, along with detailed documentation and instructions to facilitate reproducibility.

For multiple-choice QA experiments, we activated four core tools within the AI agent: `perform_comparison, enable_search, relevance_analysis`, and `locate_evidence`. Accuracy was used as the primary evaluation metric, consistent with standard practices in the field ^5,6,13,15,45^. In the open-ended QA setting, we removed predefined answer options from the prompts and extended the `generate_options` tool by building it on top of the same four tools used in the multiple-choice setting. Performance was evaluated by cosine similarity based on two state-of-the-art embedding models: `SFR-Embedding-2_R` (SFR) from Salesforce Research and `gte-Qwen2-7B-instruct` (GTE) from Alibaba Group ^36,46^. We also employed BERTScore’s F1 metric, calculated using Microsoft’s `deberta-xlarge-mnli` model, to compare the model-generated answer against ground truth ^47^. See Supplementary Sections Designed tools and Evaluation models for open-ended question answering for more details.

## RESULTS AND DISCUSSION

### Evaluation of multiple-choice benchmarks

Our agentic system achieved state-of-the-art performance across multiple-choice medical QA benchmarks, surpassing all evaluated models on USMLE Step 1, Step 2, and MedExpQA (Table 1). Specifically, it achieved 82.98% on Step 1 and 86.24% on Step 2, representing relative improvements of 2.31% and 4.57%, respectively, over GPT-4, which is the strongest baseline. On MedExpQA, where GPT-4 was not available, our model outperformed the next-best model (OpenBioLLM 70B at 71.20%) by a relative margin of 7.20%. For USMLE Step 3, our model reached 88.52%, narrowly trailing GPT-4 (89.78%) by only 1.26%. On MedQA, it scored 73.29%, which is 5.58% below GPT-4 but still ahead of all open-source models. When compared to the strongest open-source baseline, PodGPT (70B), our model demonstrated consistent and significant gains: 9.58% on Step 1, 13.76% on Step 2, 13.93% on Step 3, 8.25% on MedQA, and 15.20% on MedExpQA.

### Evaluation of open-ended medical questions

Our agentic system achieved the highest performance across all five benchmarks in the open-ended question answering setting, outperforming all baseline models on nearly every metric (Table 2). For semantic textual similarity measured using SFR model, it achieved the top score on four of five benchmarks, including USMLE Step 1 (0.87), Step 2 (0.85), Step 3 (0.86), and MedExpQA (0.84), while ranking second on MedQA (0.85 *vs.* 0.86 from OpenBioLLM 70B). While measured by the GTE model, it outperformed all baselines on USMLE Steps 1–3 (0.66, 0.62, and 0.65 respectively), and was second-best on MedQA (0.61) and MedExpQA (0.60). Similarly, our system achieved the highest or second-highest BERTScore on all benchmarks, tying for the highest score on USMLE Step 1 (0.68), Step 2 (0.67) and MedExpQA (0.65), and ranking second on USMLE Step 3 (0.70 *vs.* 0.71 from OpenBioLLM 70B) and MedQA (0.67 *vs.* 0.70 from OpenBioLLM 70B).

**Table 2.**
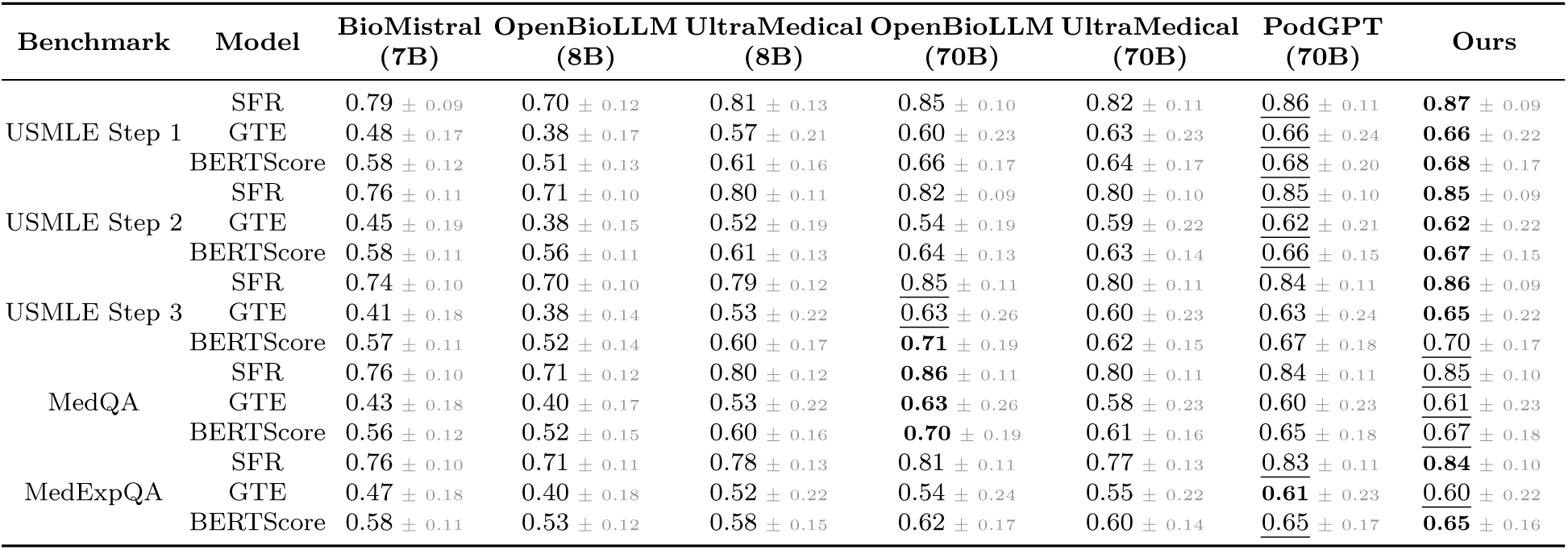
Performance evaluation on open-ended medical questions. This table reports model performance without answer choices using three embedding-based evaluation metrics: semantic textual similarity scores computed by two state-of-the-art embedding models (SFR and GTE) and BERTScore. Results are shown as mean ± standard deviation across five benchmarks (USMLE Steps 1–3, MedQA, and MedExpQA). **Bold** indicates the highest score, and <u>underlined</u> indicates the second-highest score for each metric within each benchmark.

| Benchmark | Model | BioMistral (7B) | OpenBioLLM (8B) | UltraMedical (8B) | OpenBioLLM (70B) | UltraMedical (70B) | PodGPT (70B) | Ours |
| --- | --- | --- | --- | --- | --- | --- | --- | --- |
| USMLE Step 1 | SFR | 0.79 ± 0.09 | 0.70 ± 0.12 | 0.81 ± 0.13 | 0.85 ± 0.10 | 0.82 ± 0.11 | 0.86 ± 0.11 | **0.87** ± 0.09 |
| | GTE | 0.48 ± 0.17 | 0.38 ± 0.17 | 0.57 ± 0.21 | 0.60 ± 0.23 | 0.63 ± 0.23 | 0.66 ± 0.24 | **0.66** ± 0.22 |
| | BERTScore | 0.58 ± 0.12 | 0.51 ± 0.13 | 0.61 ± 0.16 | 0.66 ± 0.17 | 0.64 ± 0.17 | 0.68 ± 0.20 | **0.68** ± 0.17 |
| USMLE Step 2 | SFR | 0.76 ± 0.11 | 0.71 ± 0.10 | 0.80 ± 0.11 | 0.82 ± 0.09 | 0.80 ± 0.10 | 0.85 ± 0.10 | **0.85** ± 0.09 |
| | GTE | 0.45 ± 0.19 | 0.38 ± 0.15 | 0.52 ± 0.19 | 0.54 ± 0.19 | 0.59 ± 0.22 | 0.62 ± 0.21 | **0.62** ± 0.22 |
| | BERTScore | 0.58 ± 0.11 | 0.56 ± 0.11 | 0.61 ± 0.13 | 0.64 ± 0.13 | 0.63 ± 0.14 | 0.66 ± 0.15 | **0.67** ± 0.15 |
| USMLE Step 3 | SFR | 0.74 ± 0.10 | 0.70 ± 0.10 | 0.79 ± 0.12 | 0.85 ± 0.11 | 0.80 ± 0.11 | 0.84 ± 0.11 | **0.86** ± 0.09 |
| | GTE | 0.41 ± 0.18 | 0.38 ± 0.14 | 0.53 ± 0.22 | 0.63 ± 0.26 | 0.60 ± 0.23 | 0.63 ± 0.24 | **0.65** ± 0.22 |
| | BERTScore | 0.57 ± 0.11 | 0.52 ± 0.14 | 0.60 ± 0.17 | 0.71 ± 0.19 | 0.62 ± 0.15 | 0.67 ± 0.18 | 0.70 ± 0.17 |
| MedQA | SFR | 0.76 ± 0.10 | 0.71 ± 0.12 | 0.80 ± 0.12 | 0.86 ± 0.11 | 0.80 ± 0.11 | 0.84 ± 0.11 | 0.85 ± 0.10 |
| | GTE | 0.43 ± 0.18 | 0.40 ± 0.17 | 0.53 ± 0.22 | 0.63 ± 0.26 | 0.58 ± 0.23 | 0.60 ± 0.23 | 0.61 ± 0.23 |
| | BERTScore | 0.56 ± 0.12 | 0.52 ± 0.15 | 0.60 ± 0.16 | 0.70 ± 0.19 | 0.61 ± 0.16 | 0.65 ± 0.18 | 0.67 ± 0.18 |
| MedExpQA | SFR | 0.76 ± 0.10 | 0.71 ± 0.11 | 0.78 ± 0.13 | 0.81 ± 0.11 | 0.77 ± 0.13 | 0.83 ± 0.11 | **0.84** ± 0.10 |
| | GTE | 0.47 ± 0.18 | 0.40 ± 0.18 | 0.52 ± 0.22 | 0.54 ± 0.24 | 0.55 ± 0.22 | 0.61 ± 0.23 | 0.60 ± 0.22 |
| | BERTScore | 0.58 ± 0.11 | 0.53 ± 0.12 | 0.58 ± 0.15 | 0.62 ± 0.17 | 0.60 ± 0.14 | 0.65 ± 0.17 | **0.65** ± 0.16 |

### Analysis of tool usage

Tool usage patterns revealed that the agent adapted its strategy to the complexity of each benchmark (Fig. 2a & Fig. 2b). While `perform_comparison` remained a consistent first-line tool across all exams, `enable_search` was used selectively, indicating the agent’s discretion in deciding when external evidence was necessary to resolve clinical uncertainty. The progressively higher use of `relevance_analysis` and `locate_evidence` tools from Step 1 to Step 3 underscores the agent’s increasing reliance on iterative evidence appraisal and grounding in more advanced clinical scenarios. This aligns with the expectation that Step 3 questions, which often involve multi-system reasoning or longitudinal management, demand a deeper chain-of-thought and external validation. The wide distribution in the number of calls to these tools further supports the hypothesis that the agent’s behavior is not hardcoded but context-dependent. In particular, questions that required repeated invocations of `relevance_analysis` and `locate_evidence` likely reflected either ambiguous clinical presentations or sparse initial document matches, prompting further rounds of evidence screening. Such behavior demonstrates the value of the cache-and-prune memory mechanism, which allowed the agent to incrementally accumulate, filter, and retain salient information while pruning irrelevant context. This architecture enabled scalable reasoning over long contexts without overwhelming the model’s input window, supporting robust performance even in highly iterative diagnostic tasks. Overall, the tool usage patterns validate both the flexibility and compositional reasoning capabilities of the agent in adapting to a diverse range of clinical question formats.

**Figure 2.**
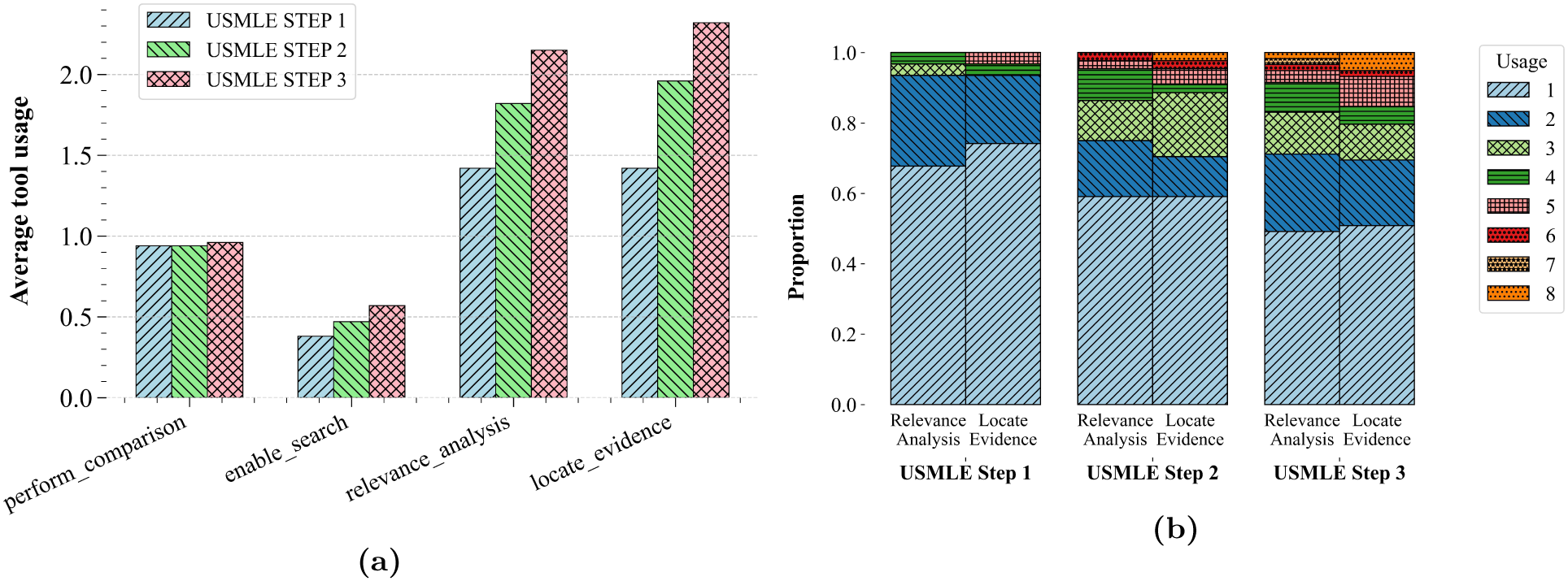
Tool usage statistics across USMLE benchmarks. (a) Bar plot showing the average number of times each tool was invoked per question across the USMLE Step 1, Step 2, and Step 3 benchmarks. Tools include `perform_comparison, enable_search, relevance_analysis`, and `locate_evidence`. (b) Stacked bar plot indicating the proportion of tool usage frequencies (from 1 to 8 calls) for `relevance_analysis` and `locate_evidence`, grouped by USMLE exam.

### Ablation studies

We compared performance with and without tool access to evaluate the impact of incorporating tools into the agentic pipeline. Specifically, we performed evaluation using structured instructions *I* without tool access (w/o Tools), and using the same instructions with full access to the toolset *T* (Ours). As shown in Table 3, tool integration led to performance improvements: 1.07% on USMLE Step 1, 3.67% on USMLE Step 2, and 4.91% on Step 3, with an average gain of 3.22% across all of them. These results underscore the value of equipping the agent with specialized tools.

**Table 3.** Impact of core components of the agentic system. Performance comparison of the agentic system with ablated versions lacking key components: tool integration, cache-and-prune memory mechanism, and external evidence search. Values for ablations indicate the relative percentage drop in accuracy compared to the full model across USMLE Step 1, Step 2, and Step 3 benchmarks.

| Benchmark | USMLE Step 1 | USMLE Step 2 | USMLE Step 3 | Average |
| --- | --- | --- | --- | --- |
| Ours | **82.98** | **86.24** | **88.52** | **85.91** |
| w/o Tools | -1.07 | -3.67 | -4.91 | -3.22 |
| w/o Cache & Prune | -1.07 | -2.75 | -3.27 | -2.36 |
| w/o Evidence Search | -2.13 | -3.67 | -6.55 | -4.12 |

To isolate the contribution of individual components, we conducted targeted ablations. Removing the `relevance_analysis` and `locate_evidence` tools (denoted w/o Cache & Prune) resulted in an average drop of 2.36%, with performance reductions of 1.07%, 2.75%, 3.27% on USMLE Step 1-3, highlighting the utility of the iterative memory mechanism. When we removed the `enable_search` tool and the document retrieval and reranking modules (`w/o Evidence Search`), performance dropped by 4.12% on average, with declines of 2.13%, 3.67%, and 6.55% on Steps 1, 2, and 3, respectively, emphasizing the critical role of external evidence in clinical reasoning.

We evaluated how the number of documents retrieved and reranked influenced the performance (Figure 3). Accuracy generally improved with increasing context length up to TopR = 32, beyond which gains plateaued. For Step 2, performance peaked at TopR = 8 with a 7.80% improvement over GPT-4 and remained stable (5.60% gain) from TopR = 32 onward. Step 1 exhibited a similar trend, with gains peaking at 5.50% at TopR = 4 and plateauing beyond TopR = 8. In contrast, while step 3 exhibited lower performance relative to GPT-4, its performance fluctuated slightly at lower TopR values and stabilized around −1.40% to −0.50% from TopR = 4 onward. These results highlight the effectiveness of our cache-and-prune memory bank in leveraging extended context efficiently, while also demonstrating the diminishing utility of low-ranked evidence beyond TopR = 32.

**Figure 3.**
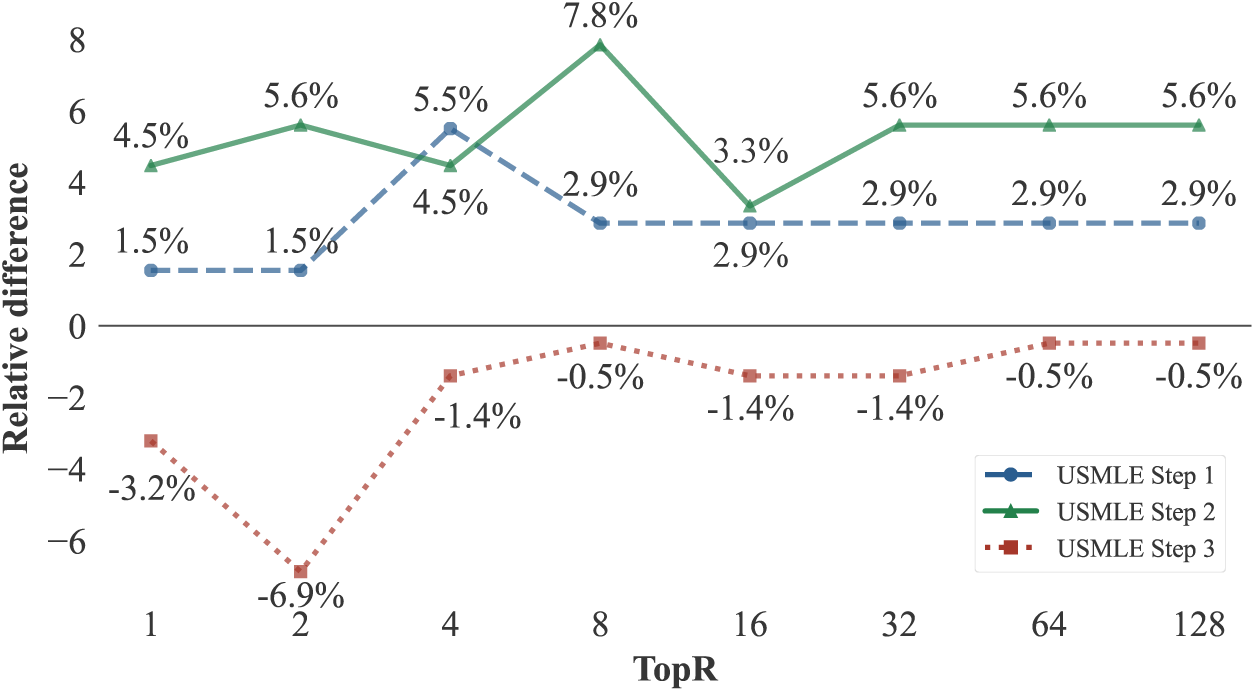
Impact of evidence context length. The figure shows the relative performance change on USMLE Step 1, Step 2, and Step 3 benchmarks as a function of the number of top reranked documents (TopR) processed by the agentic system. Each point represents the performance difference relative to GPT-4. Different line styles and colors indicate the benchmark type. The y-axis shows the relative difference in accuracy, and the x-axis denotes the number of retrieved documents.

### Limitations and future work

Despite the strong performance of our agentic system, some limitations highlight important directions for future research. First, while our system is designed as a general-purpose medical QA agent, its toolset may require domain-specific customization to handle specialized tasks, such as rare disease diagnosis or surgical decision-making. Incorporating adaptive or plug-and-play tools tailored to niche clinical domains could expand its applicability. Second, the sequential execution of tools, particularly for evidence retrieval and analysis, can introduce latency and limit scalability in real-time or high-throughput settings. Future work will explore parallelized tool execution, caching strategies across sessions, and learned policies for tool invocation to improve computational efficiency. Third, while our evaluation covered a range of benchmarks, real-world clinical scenarios often involve ambiguous, noisy or incomplete data. Expanding evaluations to include complex settings such as NEJM clinicopathological conferences, longitudinal case reports, or multimodal inputs will be important to assess robustness in high-stakes use cases ^1,48^.

Looking ahead, we envision broader societal impacts of our work in democratizing medical expertise through accessible, open-source AI systems. However, these benefits must be pursued alongside safeguards for transparency, accountability, and patient safety. As tool-based agents become more capable, interdisciplinary collaboration between clinicians, ethicists, and technologists will be important to ensure their responsible integration into clinical workflows.

## CONCLUSION

Our study advances the application of LLMs in medicine through an open-source agent that orchestrates retrieval, evidence refinement, and reasoned diagnosis in a cohesive workflow. By leveraging adaptive tools for on-demand processing and a memory system that selectively preserves critical insights, we enable more fluid and context-aware analysis, sidestepping common pitfalls like fixed prompts and truncated inputs. Our evaluations on diverse QA datasets reveal consistent advantages over established baselines, with marked improvements in precision for foundational and applied medical knowledge. Overall, this framework enhances diagnostic fidelity, accessibility, and adaptability through tool-based reasoning, laying a foundation for reliable, scalable AI that aligns with evolving healthcare demands and enables patient-centered innovations.

## Data Availability

All data produced in the present study are available upon reasonable request to the authors.

## ACKNOWLEDGMENTS

This project was supported by grants from the National Institute on Aging’s Artificial Intelligence and Technology Collaboratories (P30-AG073104 & P30-AG073105), and the National Institutes of Health (R01-NS142076, R01-HL159620, R01-AG062109, and R01-AG083735).

## DATA AND CODE AVAILABILITY

The clinical case data from NEJM used in this study are not publicly available and can be obtained under an exclusive licensing agreement with the NEJM Group. All other datasets used in this work, sourced from publicly accessible platforms such as PubMed Central, ClinicalTrials.gov, and the National Library of Medicine, will be released via Hugging Face (https://huggingface.co/vkola-lab) under a CC-BY-NC-ND license. The full source code developed for this study, including all implementation and evaluation scripts, will be made publicly available on GitHub (https://github.com/vkola-lab), along with detailed documentation and instructions to facilitate reproducibility.

