## Supplement for "Agentic memory-augmented retrieval and evidence grounding for medical question-answering tasks"

### Database for evidence retrieval

We constructed a comprehensive retrieval-augmented generation (RAG) evidence corpus by aggregating content from six trusted medical and scientific sources to ensure clinical relevance, diversity, and open accessibility. A summary of the dataset statistics, including the number of segmented snippets and their average token lengths (computed using the `Qwen2.5-72B-Instruct` tokenizer), is provided in Table 1. The corpus includes research articles published under Creative Commons licenses from leading biomedical journals indexed in PubMed Central, with specific journal titles and article counts detailed in Table 2[1]. We also incorporated clinical trial records from ClinicalTrials.gov, filtering for studies that had completed recruitment and were classified as Phase 3 or Phase 4, or that investigated device-based or behavioral interventions[2]. This selection yielded 156,887 trials as of March 2025. To enhance real-world clinical applicability, we included 1,479 clinical case reports published by the *New England Journal of Medicine* between 2016 and March 2025. We further adopted pre-indexed corpora of PubMed abstracts and Wikipedia entries from Xiong et al.[3], which have demonstrated strong utility for medical question answering tasks. Finally, we leveraged 8,226 open-access medical textbooks from the NLM LitArch Open Access Subset, hosted by the U.S. National Library of Medicine[4]. Together, these six sources form the backbone of our evidence retrieval module, supporting the agent's multi-step diagnostic reasoning with high-quality, domain-relevant content.

**Table 1. Overview of data sources for evidence retrieval.** This table summarizes the six corpora comprising our RAG database. For each source, we report the number of full documents, the number of tokenized text snippets used for retrieval, and the average token length per document (as computed using the `Qwen2.5-72B-Instruct` tokenizer). Databases are listed in descending order of document count.

| Corpus | Number of Docs | Number of Snippets | Average Length |
|---|---|---|---|
| PubMed Abstracts | 23,897,881 | 23,897,881 | 290.01 |
| Wikipedia | 6,458,670 | 29,642,311 | 166.47 |
| Clinical Trials | 156,887 | 4,177,121 | 268.33 |
| PubMed Central Articles | 123,194 | 8,155,929 | 202.46 |
| Textbooks | 8,226 | 2,224,013 | 207.95 |
| Clinical Cases | 1,479 | 17,821 | 215.61 |

**Table 2. Journals and article counts included from PubMed Central.** This table lists the 74 most represented journals in our corpus, sorted in descending order by article count. These journals span general medicine, specialty domains, and global health, contributing to a diverse and comprehensive retrieval corpus. The final row reports the total number of included articles from all journals.

| Journal Title | Article Count | Journal Title | Article Count |
|---|---|---|---|
| BMJ Open | 37,488 | JAMA Ophthalmol | 434 |
| Proc Natl Acad Sci U S A | 16,619 | Lancet HIV | 387 |
| JAMA Netw Open | 10,824 | BMJ Health Care Inform | 366 |
| Nature | 8,148 | JAMA Surg | 287 |
| Cell | 4,811 | BMJ Neurol Open | 282 |
| Science | 4,660 | JAMA Dermatol | 279 |
| BMJ | 3,636 | Lancet Psychiatry | 270 |
| BMJ Glob Health | 3,460 | Lancet Public Health | 264 |
| N Engl J Med | 2,159 | BMJ Support Palliat Care | 262 |
| BMJ Open Qual | 1,569 | BMJ Nutr Prev Health | 254 |
| JAMA | 1,552 | Lancet Respir Med | 252 |
| BMJ Open Diabetes Res Care | 1,434 | JAMA Cardiol | 239 |
| Lancet | 1,344 | Lancet Diabetes Endocrinol | 225 |
| Neurology | 1,216 | Lancet Microbe | 167 |
| BMJ Open Sport Exerc Med | 1,201 | BMJ Ment Health | 167 |
| Lancet Reg Health West Pac | 1,196 | JAMA Otolaryngol Head Neck Surg | 164 |
| BMJ Case Rep | 1,190 | Lancet Planet Health | 162 |
| BMJ Paediatr Open | 1,145 | Lancet Haematol | 157 |
| Lancet Reg Health Eur | 1,077 | BMJ Med | 154 |
| BMJ Open Respir Res | 1,031 | Lancet Child Adolesc Health | 154 |
| Lancet Reg Health Am | 901 | BMJ Evid Based Med | 136 |
| Ann Intern Med | 881 | Lancet Digit Health | 124 |
| Lancet Glob Health | 805 | BMJ Surg Interv Health Technol | 120 |
| JAMA Intern Med | 797 | Lancet Gastroenterol Hepatol | 117 |
| Lancet Infect Dis | 676 | BMJ Oncol | 114 |
| BMJ Open Ophthalmol | 656 | Lancet Healthy Longev | 102 |
| JAMA Neurol | 639 | BMJ Sex Reprod Health | 100 |
| JAMA Health Forum | 628 | BMJ Mil Health | 64 |
| BMJ Open Gastroenterol | 625 | Lancet Rheumatol | 61 |
| Lancet Oncol | 613 | BMJ Open Sci | 49 |
| BMJ Qual Saf | 601 | BMJ Innov | 46 |
| JAMA Psychiatry | 597 | BMJ Simul Technol Enhanc Learn | 42 |
| JAMA Pediatr | 569 | JAMA Facial Plast Surg | 39 |
| BMJ Qual Improv Rep | 547 | Ann Intern Med Clin Cases | 6 |
| JAMA Oncol | 490 | BMJ Outcomes | 1 |
| Lancet Reg Health Southeast Asia | 464 | BMJ Clin Evid | 1 |
| BMJ Public Health | 453 | | |
| Lancet Neurol | 444 | **Total Number of Articles** | 123,194 |

**Table 3. Overview of benchmark datasets used for evaluation.** This table summarizes the five medical QA benchmarks evaluated in our study. For each dataset, we report the total number of test cases and the maximum number of answer choices presented per question.

| Benchmark | Number of Testing Cases | Number of Choices |
|---|---|---|
| USMLE Step 1[5] | 94 | 9 |
| USMLE Step 2[5] | 109 | 6 |
| USMLE Step 3[5] | 122 | 6 |
| MedQA[6] | 1,273 | 4 |
| MedExpQA[7] | 125 | 5 |

#### Experimental benchmarks

We evaluated our system using five medical question answering benchmarks: USMLE Step 1, USMLE Step 2, USMLE Step 3, MedQA, and MedExpQA (Table 3). Each benchmark includes clinical case descriptions, multiple-choice options, and a correct answer.

The USMLE is a three-step examination series designed to assess progressively advanced competencies required for medical practice in the United States. All steps primarily use multiple-choice questions structured as clinical scenarios to evaluate critical thinking and clinical judgment. Step 1 focuses on foundational knowledge in the basic sciences, including physiology, pharmacology, pathology, and disease mechanisms. It serves as a critical assessment of preclinical competencies and includes 94 clinical cases[5]. Step 2, also known as clinical knowledge, evaluates the ability to apply medical and clinical science in the context of supervised patient care. It emphasizes diagnostic reasoning, clinical management, and ethical decision-making, with a benchmark of 109 questions[5]. Step 3 assesses readiness for independent practice by testing advanced clinical reasoning and decision-making skills across complex scenarios, including diagnosis, prognosis, and patient management. This benchmark includes 122 test cases.

MedQA is a curated benchmark for four-choice, free-form medical question answering, collected after the USMLE board exams. It spans material from Steps 1 through 3 and covers a broad range of clinical knowledge and case-based scenarios. While the original dataset includes both simplified and traditional Chinese, we used the English subset, which contains $1,273$ test cases[6]. MedExpQA follows a similar format and was constructed from the Spanish national residency medical exam. It consists of 125 test cases, each with five answer choices and detailed explanations. For our evaluation, we used the translated and annotated English subset[7].

**Backbone large language models**

Our AI agent was benchmarked against closed-source and open-source models, spanning general-purpose and medical-specific LLMs. Specifically, we compared medical diagnosis performance with leading proprietary models, including OpenAI's GPT-4 and ChatGPT[8]. On the open-source front, we included recent state-of-the-art medical LLMs such as BioMistral, OpenBioLLM, UltraMedical, and PodGPT. For all the models evaluated in this study, including our AI agent, we reported performance in the zero-shot setting.

We adopted the `Qwen2.5-72B-Instruct` model as the backbone of our AI agent. The open-source Qwen series has demonstrated competitive performance against Meta's LLaMA 3.1 models on various open-domain benchmarks, including knowledge-based and math-based tasks[9]. By default, Qwen models support a context window of up to $32,768$ tokens, which can be extended to 128K tokens using the YaRN technique[10]. However, we observed a decline in instruction-following capabilities when extending the context window under vLLM version 0.6.3. Consequently, we retained the default maximum context window of $32,768$ tokens for all experiments. Due to computational resource constraints, we focused exclusively on this model as our AI agent.

GPT-4 and GPT-3.5 (ChatGPT) from OpenAI are advanced general-purpose language models that excel across a broad spectrum of real-world tasks. In the domain of medical question answering, they have achieved state-of-the-art performance and are widely regarded as strong baselines. The evaluation results for these models, specifically `gpt-4-turbo` and `gpt-3.5-turbo`, are reported in[8].

BioMistral is the first biomedical language model based on the Mistral architecture, continually pre-trained on PubMed Central articles released under Creative Commons licenses[11]. It demonstrates improved performance on medical benchmarks compared to baseline models. In our experiments, due to its $2,048$-token context window limitation, we generated up to 128 tokens. We omitted the system prompt, as the Mistral chat template did not support it.

OpenBioLLM builds upon the LLaMA 3 architecture and is available in both 8B and 70B parameter versions. These models are fine-tuned using direct preference optimization, a reinforcement learning-based alignment technique[12]. OpenBioLLM demonstrates competitive performance against both its baseline and proprietary counterparts[13]. In this study, we evaluated the 8B and 70B variants. Additionally, we configured the models with a maximum context length of $8,192$ tokens and generated up to $1,024$ tokens per response.

UltraMedical models, trained through supervised fine-tuning and preference-based learning, demonstrate competitive performance with proprietary LLMs such as OpenAI GPT-4[12,14]. In our experiments, we evaluated both the 8B model, based on LLaMA 3.1, and the 70B model, based on LLaMA 3, as the LLaMA 3.1 version of the UltraMedical 70B model was not publicly available at the time of this study.

PodGPT is a family of language models continually pre-trained on publicly available podcasts spanning the domains of science, technology, engineering, mathematics, and medicine (STEMM). Designed specifically for scientific and educational applications, these models were evaluated across a range of STEMM benchmarks, including datasets focused on medical question answering[15]. We employed the best-performing PodGPT model, based on the `Llama-3.3-70B-Instruct` architecture fine-tuned with a low-rank adapter[16]. To maintain consistency with the OpenBioLLM and UltraMedical configurations, we set the context window to $8,192$ tokens and allowed up to $1,024$ tokens to be generated.

**Table 4. Parameters used in the agent's toolset.** This table outlines the parameters and their corresponding descriptions for each tool integrated into our diagnostic framework. The tools `perform_comparison`, `enable_search`, `relevance_analysis`, and `locate_evidence` are used for multiple-choice QA tasks. For open-ended QA, the `generate_options` tool is additionally employed, generating plausible answer options for further analysis.

| Tool Name | Parameter | Parameter Description |
|---|---|---|
| `generate_options` | answers | The most likely answers based on the patient's specific condition and needs. |
| `perform_comparison` | comparisons | A structured comparison of all options, detailing their relevance to the patient's case. |
| `enable_search` | search | Answer 'yes' or 'no' to indicate search necessity. |
| `relevance_analysis` | analysis | A comprehensive analysis detailing the relevance of each document to the patient's presentation, highlighting key matches, inconsistencies, and important findings. |
| `locate_evidence` | evidence | Relevant evidence applicable to the patient's presentation, with article IDs in <quote></quote>tags. |

### Designed tools

As illustrated in Fig. 1, we designed five specialized tools to serve as the diagnostic aid kit within our AI agent. For question interpretation, the agent uses `perform_comparison` to handle multiple-choice tasks and `generate_options` for open-ended scenarios, enabling flexible reasoning formats. In particular, `generate_options` is tailored for scenarios lacking predefined choices, enabling the agent to propose plausible answer candidates from the contextual details of the case. Additionally, the `enable_search` tool determines whether external evidence is necessary to support a diagnosis. To facilitate evidence retrieval and interpretation, `relevance_analysis` evaluates the semantic alignment between the patient's case and retrieved documents, while `locate_evidence` identifies and grounds specific articles most pertinent to the diagnosis.

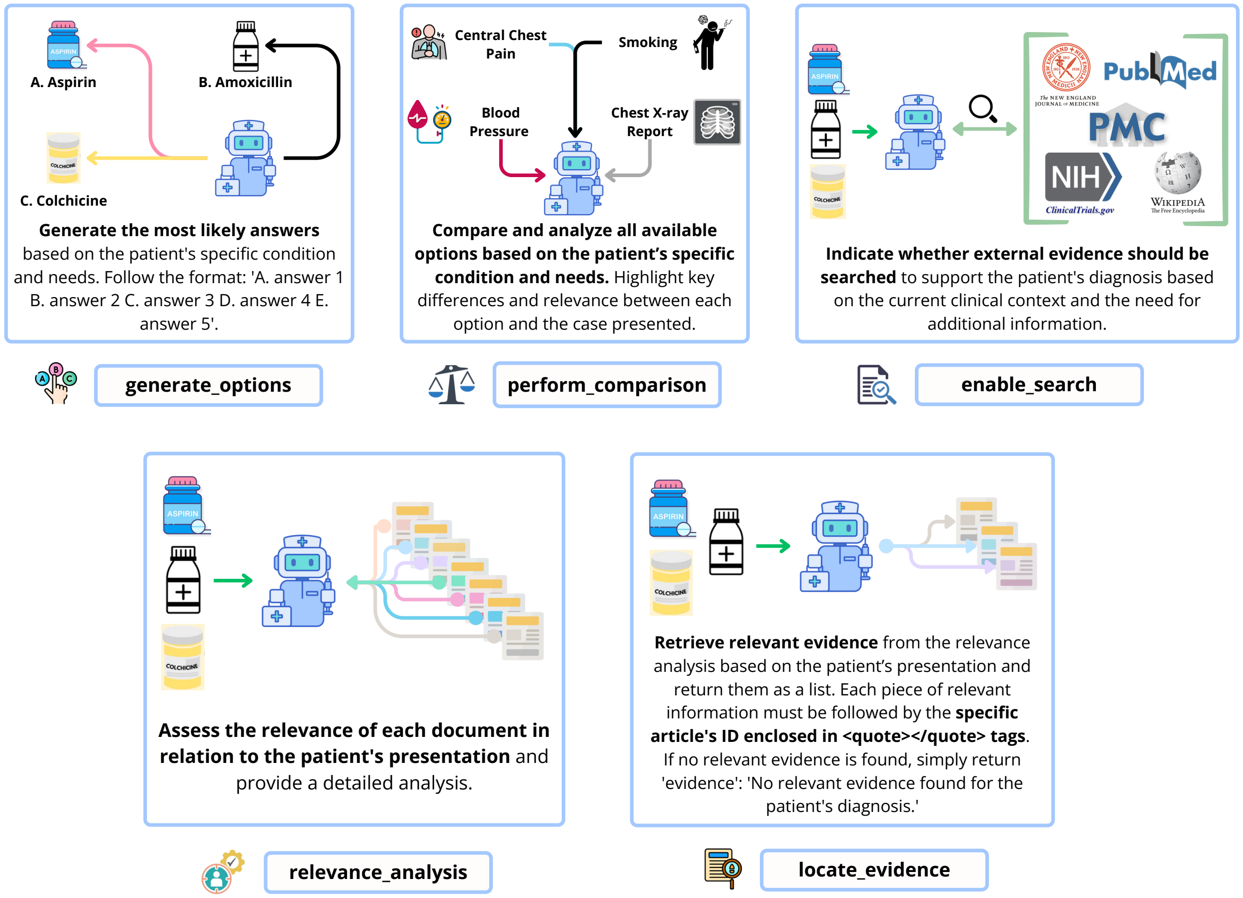


**Figure 1. Overview of specialized tools in the agentic framework.** This figure illustrates the five custom-designed tools used by the agent for medical question answering in the open-ended setting. Each tool performs a distinct role: `generate_options` first proposes potential answers to the problem, `perform_comparison` then analyzes the candidate options in the context of the problem description, `enable_search` decides whether external evidence is needed, `relevance_analysis` assesses the contextual fit of retrieved documents, and `locate_evidence` extracts grounded evidence snippets tied to article IDs. Together, these tools enable dynamic, interpretable, and evidence-grounded reasoning.

#### Evaluation models for open-ended question answering

In this work, we employed two state-of-the-art semantic similarity models, `SFR-Embedding-2_R` (SFR) and `gte-Qwen2-7B-instruct` (GTE), alongside BERTScore, enabling fine-grained semantic comparison between the model-generated responses and ground-truth answers[17–19]. For both SFR and GTE, we used the default cosine similarity to compute phrase-level similarity, while for BERTScore, we reported the F1 metric to assess alignment at the token level.

The `SFR-Embedding-2_R` model is based on the Mistral architecture with 7 billion parameters and supports input lengths of up to 4,096 tokens[17]. This model achieves strong results on the massive text embedding benchmark (MTEB), highlighting its robustness for semantic similarity tasks.

The `gte-Qwen2-7B-instruct` model was built on the Qwen2 architecture with 7 billion parameters and supports input lengths of up to 32K tokens[18]. It was instruction-tuned for a range of natural language processing tasks, including retrieval, classification, and reranking. The model ranks highly on the MTEB leaderboard, demonstrating state-of-the-art performance in semantic textual similarity.

BERTScore evaluates the semantic similarity between two phrases by computing the cosine similarity between their contextualized token embeddings, derived from a pretrained language model[19]. In our experiments, we used the `deberta-xlarge-mnli` model as the backbone for BERTScore computation. This model, with 750 million parameters, was fine-tuned on the multi-genre natural language inference tasks, making it particularly well-suited for assessing phrase and sentence-level semantic alignment in open-ended medical QA tasks.

#### Used prompts

To ensure fair and consistent evaluation, we employed a unified set of prompts across all open-source models in a direct-response format. Each model was paired with its designated chat template, as defined by its tokenizer specifications. For our AI agent, the primary prompt templates used for multiple-choice question answering are presented in Table 5, with a standardized System Prompt applied uniformly across all configurations. For the open-ended QA setting, we adapted the same templates by removing the answer choices, allowing the models to generate free-form diagnostic responses. The corresponding prompt format for open-ended questions is provided in Table 6.

**Table 5. Prompt templates for multiple-choice question answering.** This table presents the System Prompt and Prompt Template used for multiple-choice QA, along with the document formatting template and the cache-and-prune memory bank mechanism template employed by our AI agent.

| **System Prompt** |
|---|
| You are a medical professional specializing in evidence-based medicine (EBM). Your role is to answer questions using a systematic approach, integrating the best available research evidence, clinical expertise, and patient-specific factors. |
| **Prompt Template** |
| Here is the background information and question about the patient:<br><background><br>{background}<br></background><br><br>The available answer options are:<br><option><br>{option}<br></option><br><br>Follow these steps to answer the question:<br>1. Compare each option with the case details, analyzing key clues in the text to identify the best choice.<br>2. If the question can be answered through comparison, directly return the best option term with the option capital within <final_result></final_result>tags, placing the explanation outside of the <final_result>tags.<br>3. If multiple options are plausible or additional evidence is needed for better decision-making, enable search to find credible sources.<br>4. Analyze the relevance between each document and the patient's presentation, followed by a systematic search to locate relevant evidence applicable to the patient's case.<br>5. While we are continuing to provide additional evidence, iterate the previous step to analyze more additional evidence.<br>6. Once sufficient information is gathered, return the best option term with the option capital within <final_result></final_result>tags, placing the explanation outside of the <final_result>tags. |
| Document template |
| Relevant documents related to the patient's care:<br><document><br>{document}<br></document> |
| Cache-and-prune memory bank mechanism template |
| Here are the selected relevant documents related to the patient's care:<br><document><br>{document}<br></document><br><br>Review your answer and return the best option term with the option capital within the <final_result></final_result>tags, leave the explanation outside of the <final_result>tags. |

**Table 6. Prompt templates for open-ended question answering.** This table presents the Prompt Template, document formatting template, and the template for the cache-and-prune memory bank mechanism used in open-ended QA.

| **Prompt Template** |
|---|
| Here is the background information and question about the patient:<br><background><br>{background}<br></background><br><br>Follow these steps to answer the question:<br>1. Compare each option with the case details, analyzing key clues in the text to identify the best choice.<br>2. If the question can be answered through comparison, directly return the full answer term within <final_result></final_result>tags, placing the explanation outside of the <final_result>tags.<br>3. If multiple options are plausible or additional evidence is needed for better decision-making, enable search to find credible sources.<br>4. Analyze the relevance between each document and the patient's presentation, followed by a systematic search to locate relevant evidence applicable to the patient's case.<br>5. While we are continuing to provide additional evidence, iterate the previous step to analyze more additional evidence.<br>6. Once sufficient information is gathered, return the full answer term within <final_result></final_result>tags, placing the explanation outside of the <final_result>tags. |
| Document template |
| Relevant documents related to the patient's care:<br><document><br>{document}<br></document> |
| Cache-and-prune memory bank mechanism template |
| Here are the selected relevant documents related to the patient's care:<br><document><br>{document}<br></document><br><br>Review your answer and return the full answer term within the <final_result></final_result>tags, leave the explanation outside of the <final_result>tags. |

17. Meng R, Liu Y, Joty SR, Xiong C, Zhou Y, Yavuz S. SFR-Embedding-2: Advanced Text Embedding with Multi-stage Training; 2024.

18. Li Z, Zhang X, Zhang Y, Long D, Xie P, Zhang M. Towards General Text Embeddings with Multi-stage Contrastive Learning. arXiv preprint arXiv:230803281. 2023. Available from: https://doi.org/10.48550/arXiv.2308.03281.

19. Zhang T, Kishore V, Wu F, Weinberger KQ, Artzi Y. BERTScore: Evaluating Text Generation with BERT. In: The Eleventh International Conference on Learning Representations, ICLR 2020, Addis Ababa, Ethiopia, April 26-30, 2020. International Conference on Learning Representations; 2020. Available from: https://openreview.net/forum?id=SkeHuCVFDr.
